# Association of vasopressors with mortality in critically ill patients with COVID-19: A systematic review and meta-analysis

**DOI:** 10.1101/2022.05.27.22275715

**Authors:** Maria Mermiri, Georgios Mavrovounis, Eleni Laou, Nikolaos Papagiannakis, Ioannis Pantazopoulos, Athanasios Chalkias

## Abstract

**Purpose:** The effect of vasopressors on mortality of critically ill patients with COVID-19 has not been studied extensively.

**Materials and Methods:** A systematic search of PubMed, Scopus, and clinicaltrials.gov was conducted for relevant articles until January 2022. Eligibility criteria were randomized controlled and non-randomized trials. The primary outcome was mortality at latest follow-up. The quality of studies was assessed using the MINORS tool. Paired meta-analysis was used to estimate the pooled risk ratios along with their 95% Confidence Interval.

**Results:** Analyses of 21 studies (n=7900) revealed that vasopressor use is associated with mortality in patients who receive vasopressors compared to those who do not receive vasopressor therapy [RR (95%CI): 4.26 (3.15, 5.76); p<0.001]. In-hospital and 30-day mortality are significantly higher in patients who receive vasopressors [RR (95%CI): 4.60 (2.47, 8.55); p<0.001 and RR (95%CI): 2.97 (1.72, 5.14); p<0.001, respectively]. The highest mortality rate was observed with vasopressin or epinephrine, while the lowest mortality rate was observed with angiotensin-II. Also, analyses of data from 10 studies (n=3519) revealed that vasopressor use is associated with acute kidney injury [RR (95%CI): 3.17 (2.21, 4.54); p<0.001].

**Conclusion:** Vasopressor use was associated with an increase in in-hospital mortality, 30-day mortality, and acute kidney injury in critically ill patients with COVID-19.

## INTRODUCTION

Mounting evidence suggest that COVID-19 should be perceived as a new entity with its own characteristics and distinct pathophysiology, including complex immuno-inflammatory, thrombotic, and parenchymal derangements [1]. The cytokine storm and the dysregulation of host response are more severe in COVID-19-related acute respiratory distress syndrome (ARDS) than in ARDS of other causes [2–4]. SARS-CoV-2 not only infects the respiratory tract, but also injures the vascular endothelium and epithelium [5, 6].

Most critically ill patients with COVID-19 need hemodynamic support that is usually guided by the current, non-covid, surviving sepsis campaign guidelines recommending the use of vasopressors to optimize mean arterial pressure (MAP) and cardiac output and provide adequate organ perfusion [7, 8]. Most of these medications improve the hemodynamic function through enhancement of the adrenergic pathway; however, they may have important side-effects due to excessive adrenergic stimulation [9–11]. Of note, exogenous catecholamines can have a pronounced impact on inflammation and immunosuppression, metabolism, endothelial lesion, platelet activation, and coagulation [12]. As critically ill patients with COVID-19 are characterized by a similar pathophysiological substrate, exogenous vasopressors could further dysregulate their physiological cascades and aggravate outcome [13]. We therefore performed a systematic review and meta-analysis to investigate the effect of vasopressors on mortality of critically ill patients with COVID-19.

## MATERIAL AND METHODS

The protocol was registered in the PROSPERO international prospective register of systematic reviews on 13 December 2021 (CRD42021297595). This systematic review and meta-analysis was designed according to the preferred reporting items for systematic reviews and meta-analyses (PRISMA) checklist (Appendix A) [14].

### Inclusion and exclusion criteria

The inclusion criteria of the current systematic review and meta-analysis were: (1) randomized controlled trials (RCTs) and observational studies; (2) critically ill patients admitted to the intensive care (ICU) or high dependency unit (HDU), including patients admitted through the Emergency Department (ED); (3) adults (≥ 18 years old) hospitalized primarily for COVID-19; (4) SARS-CoV-2 infection confirmed by reverse transcription polymerase chain reaction test of nasopharyngeal or oropharyngeal samples; and (5) vasopressor *vs.* no vasopressor administration. We excluded animal studies, case reports, review papers, editorials, abstracts, white papers, and non-English literature. We also excluded studies about pediatric patients and non-ICU/HDU/ED patients.

### Outcomes of interest

The primary outcome was mortality at latest follow-up. Secondary outcomes was to investigate (1) the hemodynamic profiles of patients at first measuring point and after six hours [heart rate, MAP, central venous pressure (CVP), urinary output, blood lactate levels, cardiac output or cardiac index, systemic vascular resistance index, central venous oxygen saturation, oxygen delivery index, and oxygen consumption index]; (2) the number of participants who achieved the target MAP (≥65 mmHg); (3) time to achieve the target MAP; (4) adverse events including arrhythmia, acute myocardial infarction, cardiac arrest, acute mesenteric ischemia, digital ischemia, acute kidney injury (AKI); (5) vasopressor-free days; (6) ICU or HDU length of stay; (7) duration of mechanical ventilation; (8) ventilator free days; (9) hospital length of stay; and (10) all-cause mortality at 90-days.

### Search strategy

The search strategy was intended to explore all available published and unpublished studies from January 2020 to January 2022. A comprehensive initial search was employed in PubMed (MEDLINE), Scopus, and clinicaltrials.gov databases by two independent investigators (MM, GM) followed by an analysis of the textwords contained in Title/Abstract and indexed terms. A second search was conducted by combining free text words (vasopressor, epinephrine, norepinephrine, phenylephrine, vasopressin, dopamine, angiotensin-II, covid-19, critically ill, intensive care) and indexed terms with Boolean operators. Finally, a third search was conducted with the reference lists of all identified reports and articles for additional studies. Appendix B presents the exact search algorithm used for all databases.

### Data extraction

The data from each study were extracted by two independent authors (MM, GM) with a customized format. Any disagreements between the two independent authors were resolved by four other authors (EL, IP, NP, AC). Publication details (authors, year), study information (design, population, department of admission, follow-up, inclusion-exclusion criteria, number of cases/cohort-size, and subgroups), hemodynamic profile (heart rate, MAP, CVP, urinary output, blood lactate levels, cardiac output or cardiac index, systemic vascular resistance index, central venous oxygen saturation, oxygen delivery index, oxygen consumption index) at first measuring point and six hours after vasopressor use, the number of participants who achieved the target MAP and time to achieve the target MAP, adverse events, vasopressor-free days, ICU length of stay, hospital length of stay, duration of mechanical ventilation, ventilator-free days, all-cause mortality in all groups at 28 or 30 days, and all-cause mortality at 90 days were extracted in a pre-designed excel spreadsheet. The definition used for AKI and the mortality follow-up timepoints for each study are presented in Appendix C1. Authors of studies with missing data were contacted in an attempt to obtain relevant data.

### Assessment of methodological quality

Articles identified for retrieval were assessed by two independent authors (MM, GM) for methodological quality before inclusion in the review using standardized critical appraisal tools. The quality of the included observational studies was assessed using the MINORS tool [15], while the Risk of Bias 2.0 (RoB 2.0) tool was used for RCTs [16]. Any disagreements between the authors appraising the articles were resolved through discussion with the other authors.

### Data analysis and Synthesis

A paired meta-analysis was used to estimate the pooled risk ratios (RR) along with their 95% Confidence Interval (95% CI). Based on the presence of statistical heterogeneity, the meta-analysis was conducted according to fixed- or random effect models. The statistical heterogeneity was estimated by the use of the Cochran’s Q and *I*^2^ indices. When *I*^2^>50% and/or P_Q_<0.10, the random effects model was used, otherwise the fixed effects model was implemented [17]. Funnel plots as well as the Begg’s test were used to determine the existence of publication bias [18, 19]. The statistical significance was set at *p*<0.05. All statistical analyses were performed in Review Manager (Rev-Man) [Computer program], Version 5.3. Copenhagen: The Nordic Cochrane Centre, The Cochrane Collaboration, 2014.

## RESULTS

Altogether, 809 relevant citations were identified and screened, while 87 studies were included in our final assessment for possible data extraction (Fig. 1). In total, data extraction was possible in 33 studies [20–52].

**Fig. 1.**
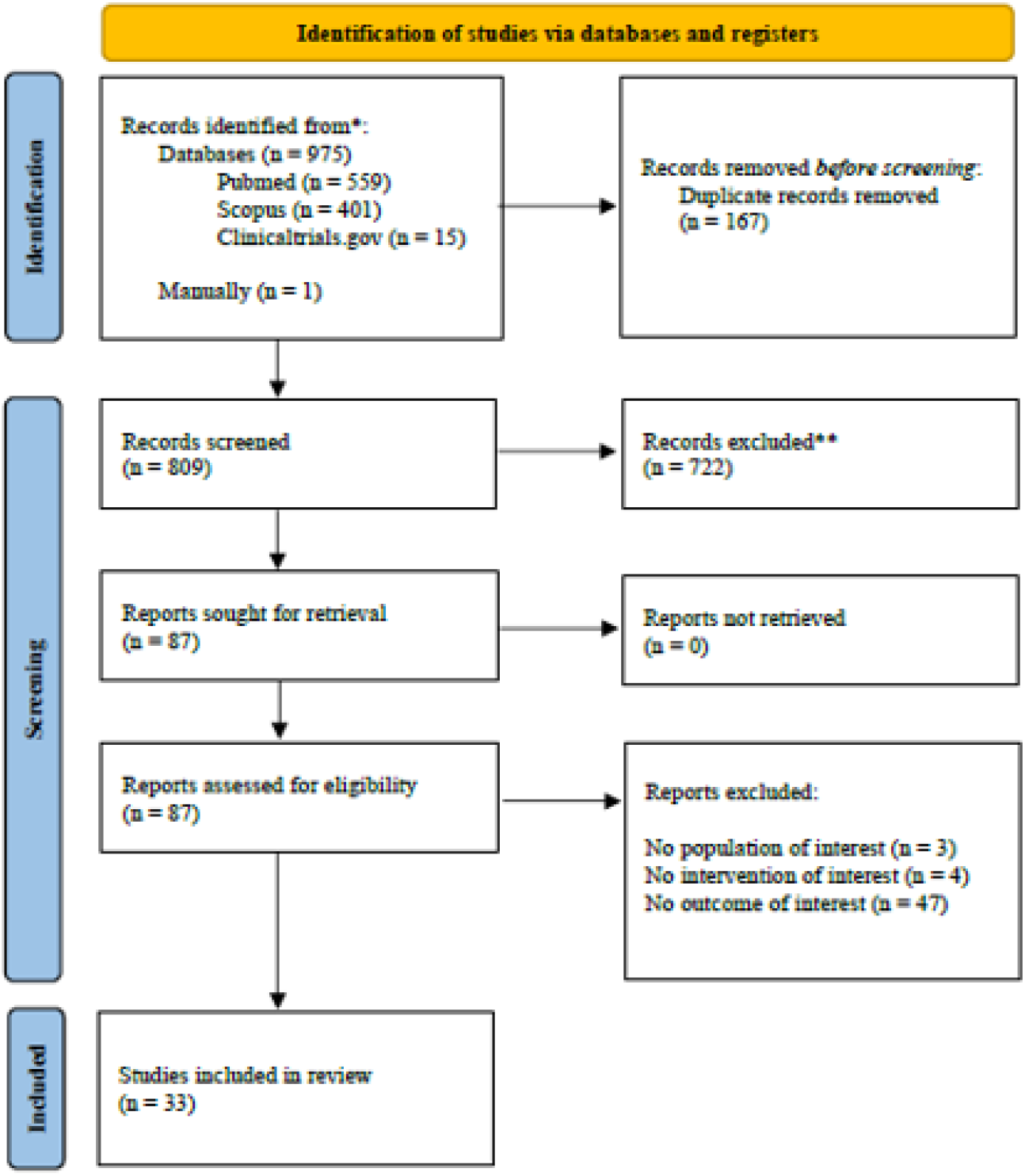
Preferred Reporting Items for Systematic Reviews and Meta-Analyses (PRISMA) diagram

### Study characteristics

All the 33 included studies were observational in their design [20–52]. The studies originated from multiple countries [20,21,23,32–35,38,42,43,45,48,51]. Nineteen studies included only patients admitted to the ICU [20,22,24,25,27,30-32,35–39,43–47,52], five studies included patients admitted to a COVID-19-dedicated HDU [28,29,33,34,42], eight studies included patients who were admitted to both HDU and ICU [21,23,26,40,41,48,49,51], and one study included Emergency Department patients who were later admitted either to the HDU of ICU [50]. Thirty studies included data about patients who received *vs.* patients who did not receive vasopressors [20-37,39-48,50,52] and were included in review. Moreover, three studies included patients who received angiotensin-II [38,49,51] and, out of those, two compared the use of angiotensin-II with other vasopressors [49, 51] (Appendix C2). Table 1 summarizes the main characteristics of the included studies.

**Table 1.**
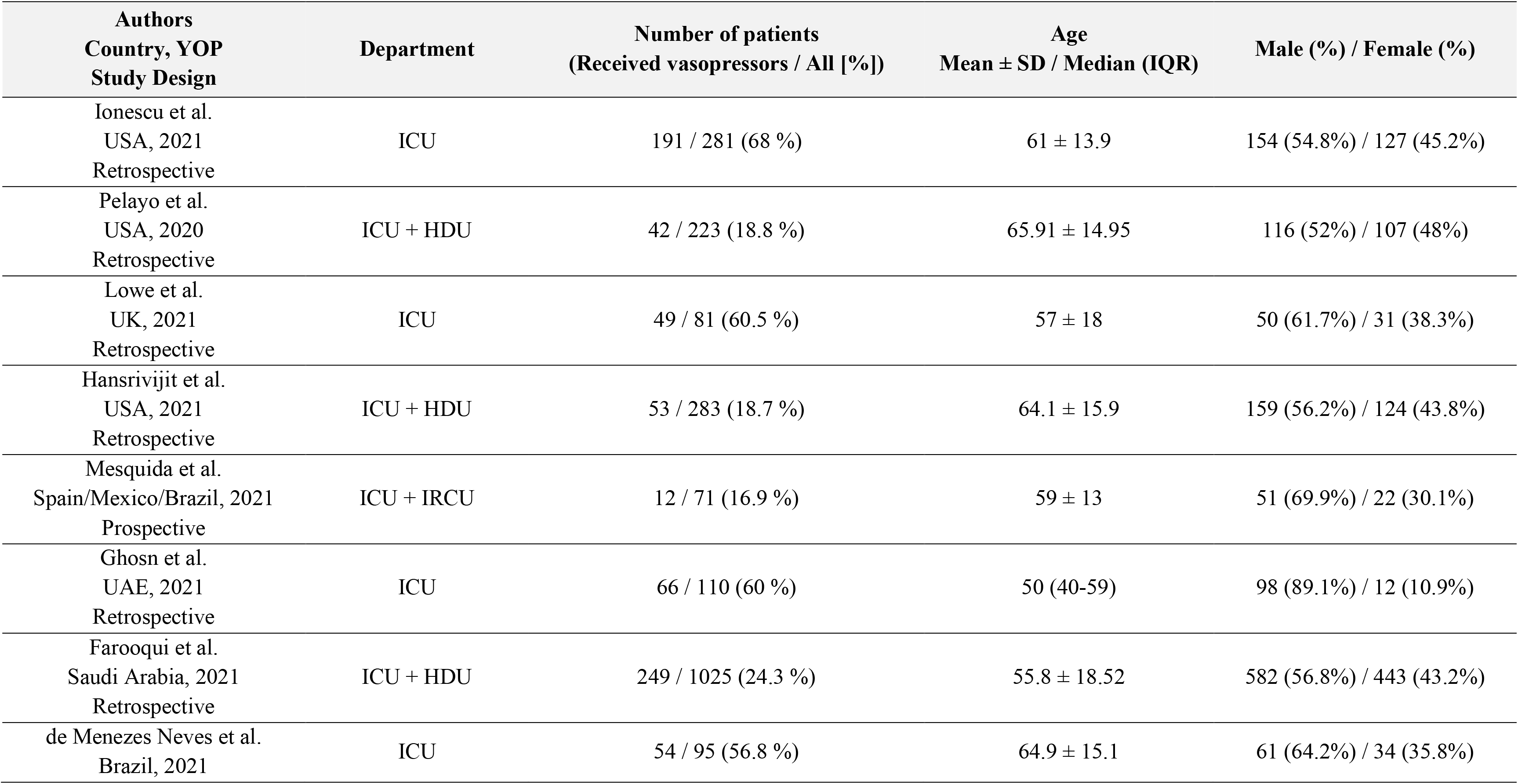
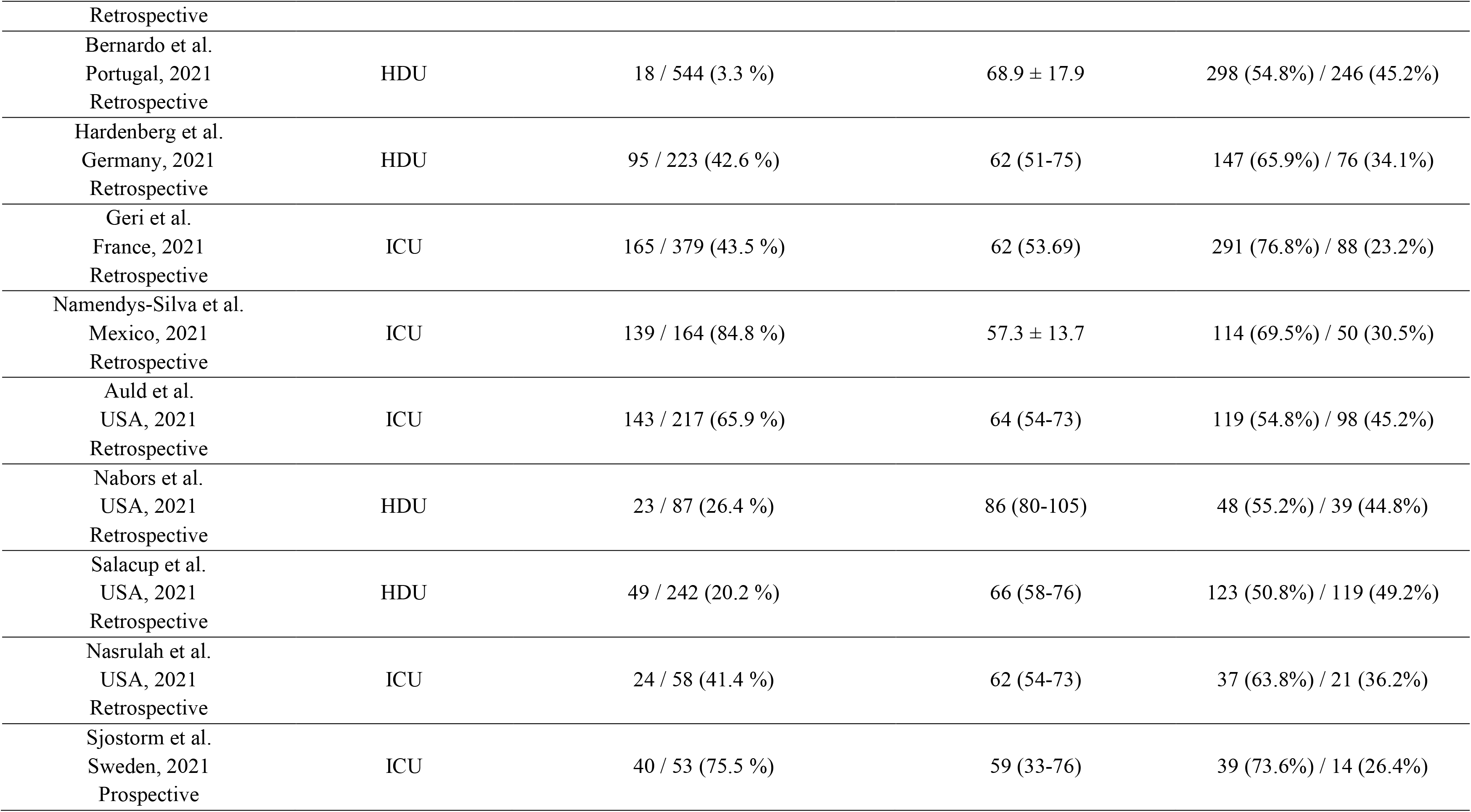
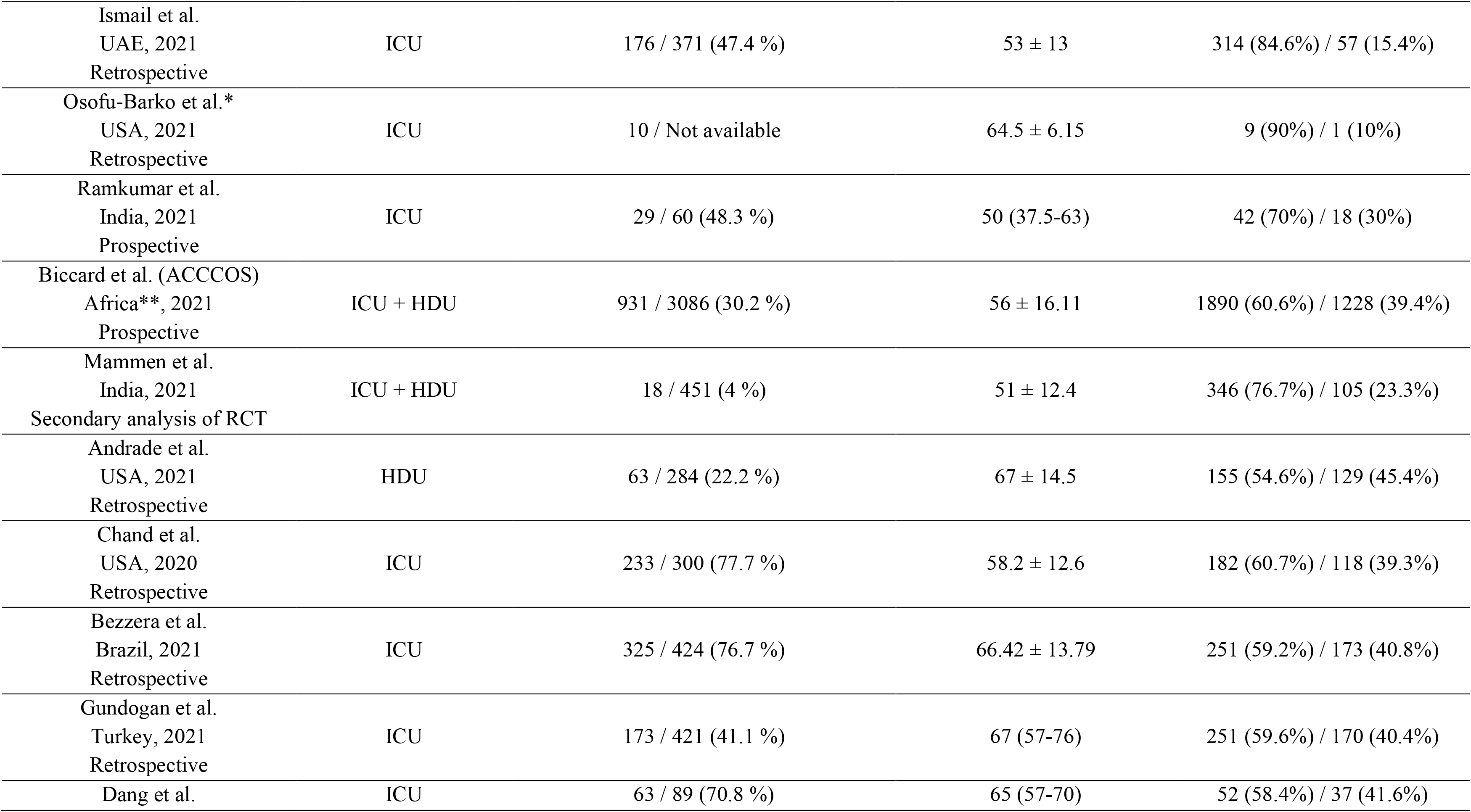
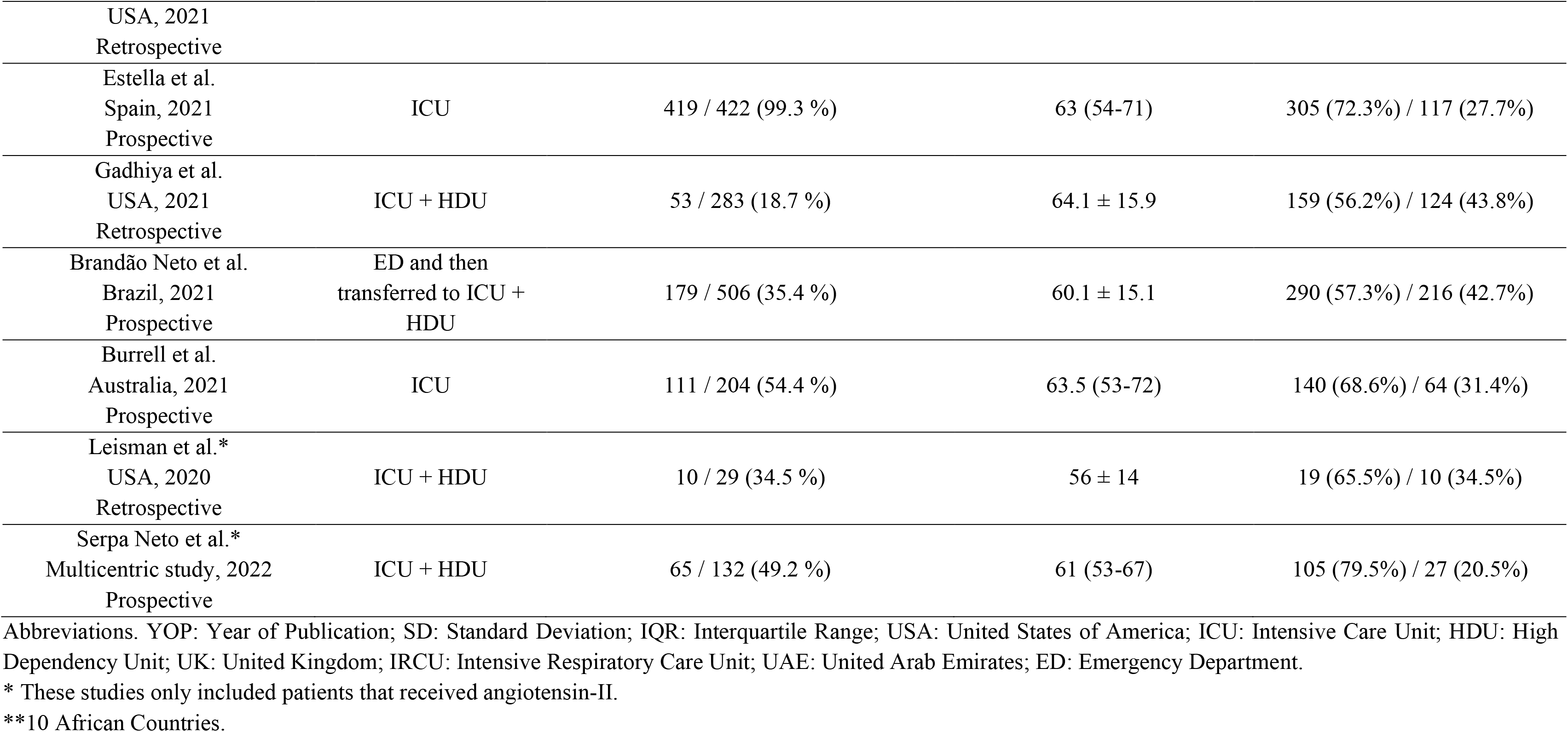
Main characteristics of the included studies.

### Synthesis including all patients

#### Primary outcome

Twenty-one out of the 33 included studies provided data on hospital mortality in patients who received *vs.* patients who did not receive vasopressors, resulting in a total population of 7900 individuals [25,27,31–37,39–48,50,52]. Due to high heterogeneity (I^2^: 94%, P_Q_<0.001), the random-effects model was implemented. Vasopressor use is associated with mortality in patients who receive vasopressors compared to those who do not receive vasopressor therapy [RR (95%CI): 4.26 (3.15, 5.76); p<0.001] (Figure 2). Visual inspection of the funnel plot (Appendix D1) and Begg’s test (p=0.86) did not reveal significant publication bias.

**Fig. 2.**
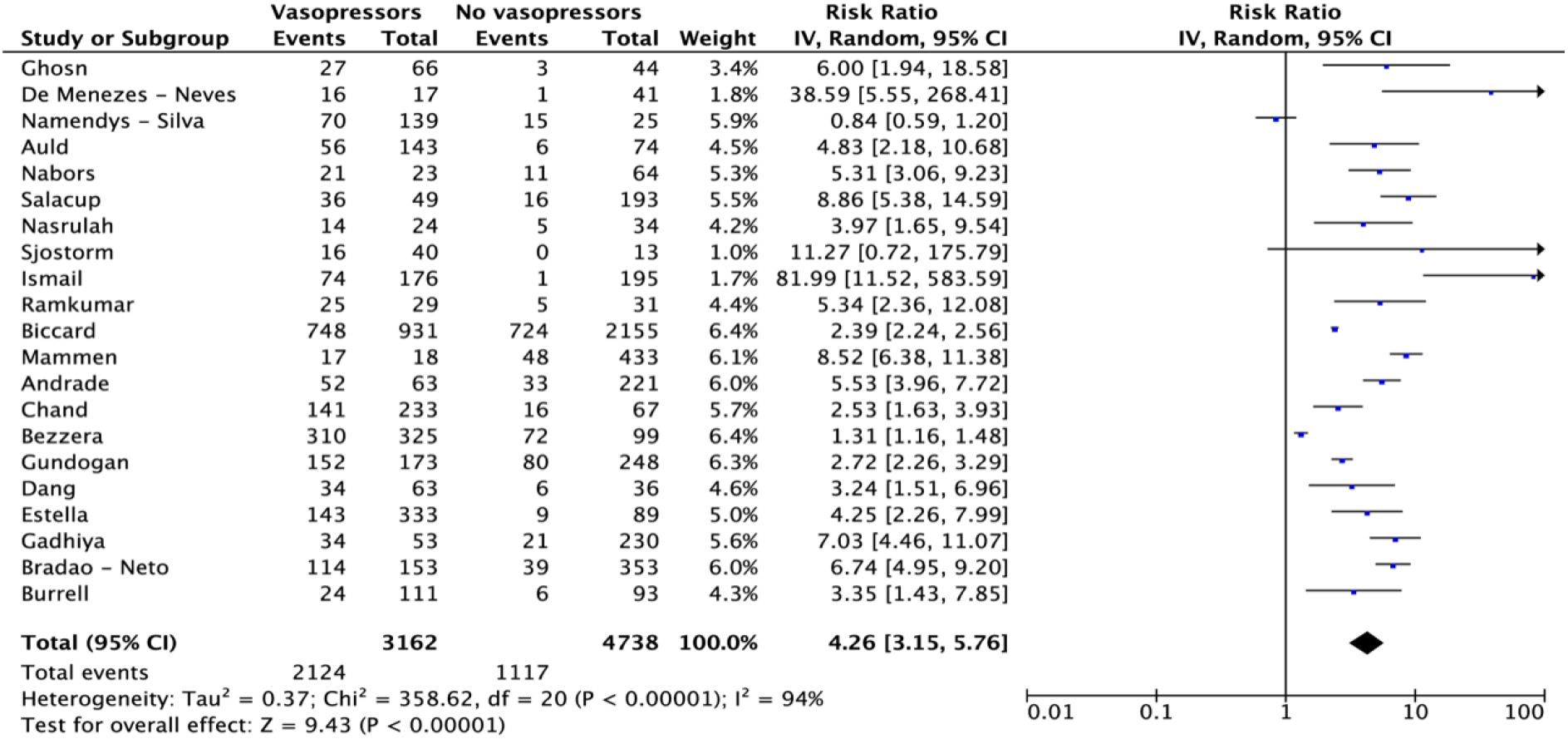
Effect of vasopressor use on mortality of critically ill patients with COVID-19

We performed subgroup analyses based on the department of admission. The results for all three subgroups, namely ICU [RR (95%CI): 3.45 (2.32, 5.13); p < 0.001], HDU [RR (95%CI): 6.25 (4.63, 8.44); p < 0.001], and ICU + HDU [RR (95%CI): 5.52 (2.51, 12.15); p < 0.001], remained statistically significant for higher mortality rates in patients who received vasopressors.

Subgroup analyses were also performed based on the mortality follow-up timepoints. Only the in-hospital and 30-day mortality subgroups had three or more studies that allowed data extraction and analysis. The in-hospital and 30-day mortality were statistically significantly higher in patients who received vasopressors [RR (95%CI): 4.60 (2.47, 8.55); p<0.001 and RR (95%CI): 2.97 (1.72, 5.14); p<0.001, respectively].

##### Effect of major vasopressors on mortality

Four studies provided data on mortality based on the specific vasopressor(s) administered [38,43,49,51]. The highest mortality rate was observed in patients treated with vasopressin or epinephrine (78% and 76%, respectively) [43]. Three of those studies investigated the role of angiotensin-II as a sole or second-, third-, fourth-, or fifth-line vasopressor agent [38,49,51]. These studies showed the lowest mortality rate. The relevant data are depicted in Table 2.

**Table 2.** Data on mortality based on major vasopressors

| **Authors**<br>**Country, YOP**<br>**Study Design** | **Intervention Group: Deaths / All (%)** | **Comparator Group: Deaths / All (%)** |
| --- | --- | --- |
| Chand et al.<br>USA, 2020<br>Retrospective | Any vasopressor support: 141 / 233 (61%)<br>Norepinephrine ± other vasopressors: 138 / 226 (61%)<br>Phenylephrine ± other vasopressors: 53 / 89 (60%)<br>Vasopressin ± other vasopressors: 81 / 104 (78%)<br>Epinephrine ± other vasopressors: 19 / 25 (76%) | No vasopressor support: 16 / 67 (24%) |
| Leisman et al.<br>USA, 2020<br>Retrospective | Angiotensin-II ± other vasopressors: 4 / 10 (40%) | Other vasopressors: 10 / 19 (53%) |
| Serpa Neto et al.<br>Multicentric study, 2022<br>Prospective | Angiotensin-II ± norepinephrine: 35 / 65 (54%) | Other vasopressors: 27 / 67 (40%) |
| Ofosu-Barko et al.<br>USA, 2021<br>Retrospective | Angiotensin-II + other vasopressors: 4 / 10 (40%) | - |

#### Secondary outcomes

##### Acute Kidney Injury

Ten studies provided data on AKI in patients who received *vs.* patients who did not receive vasopressors, resulting in a total population of 3519 individuals [21-23,25-30,44]. Due to high heterogeneity (I^2^: 92%, P_Q_<0.001), the random-effects model was implemented. In addition, vasopressor use is associated with AKI in patients who receive vasopressors compared to those who do not receive vasopressor therapy [RR (95%CI): 3.17 (2.21, 4.54); p<0.001] (Figure 3).

**Fig. 3.**
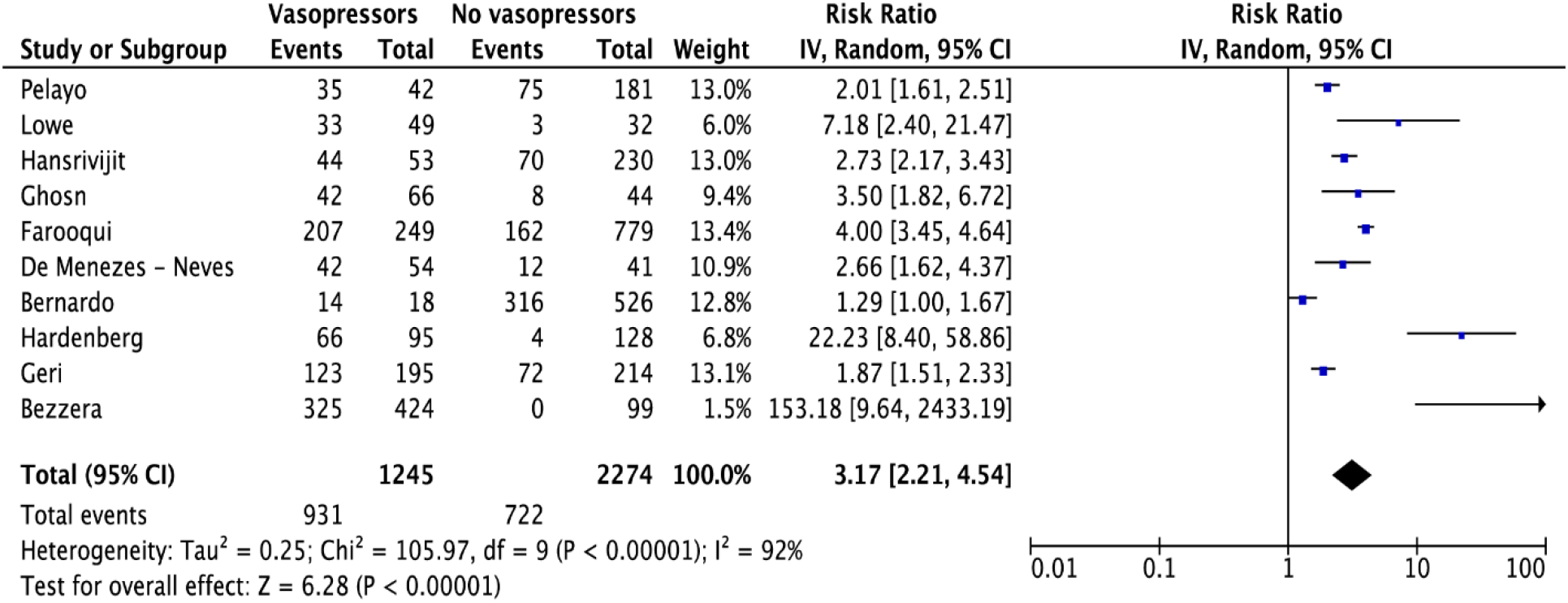
Effect of vasopressor use on the incidence of acute kidney injury

Subgroup analyses were performed based on the definition of AKI that was used in the included studies. Only the subgroup with patients at all KDINGO stages included more than three studies, allowing for meta-analysis to be performed. Specifically, vasopressor use is associated with AKI in patients who receive vasopressors compared to those who do not receive vasopressor therapy [RR (95%CI): 2.29 (1.67-3.14); p < 0.001] (Figure 4).

**Fig. 4.**
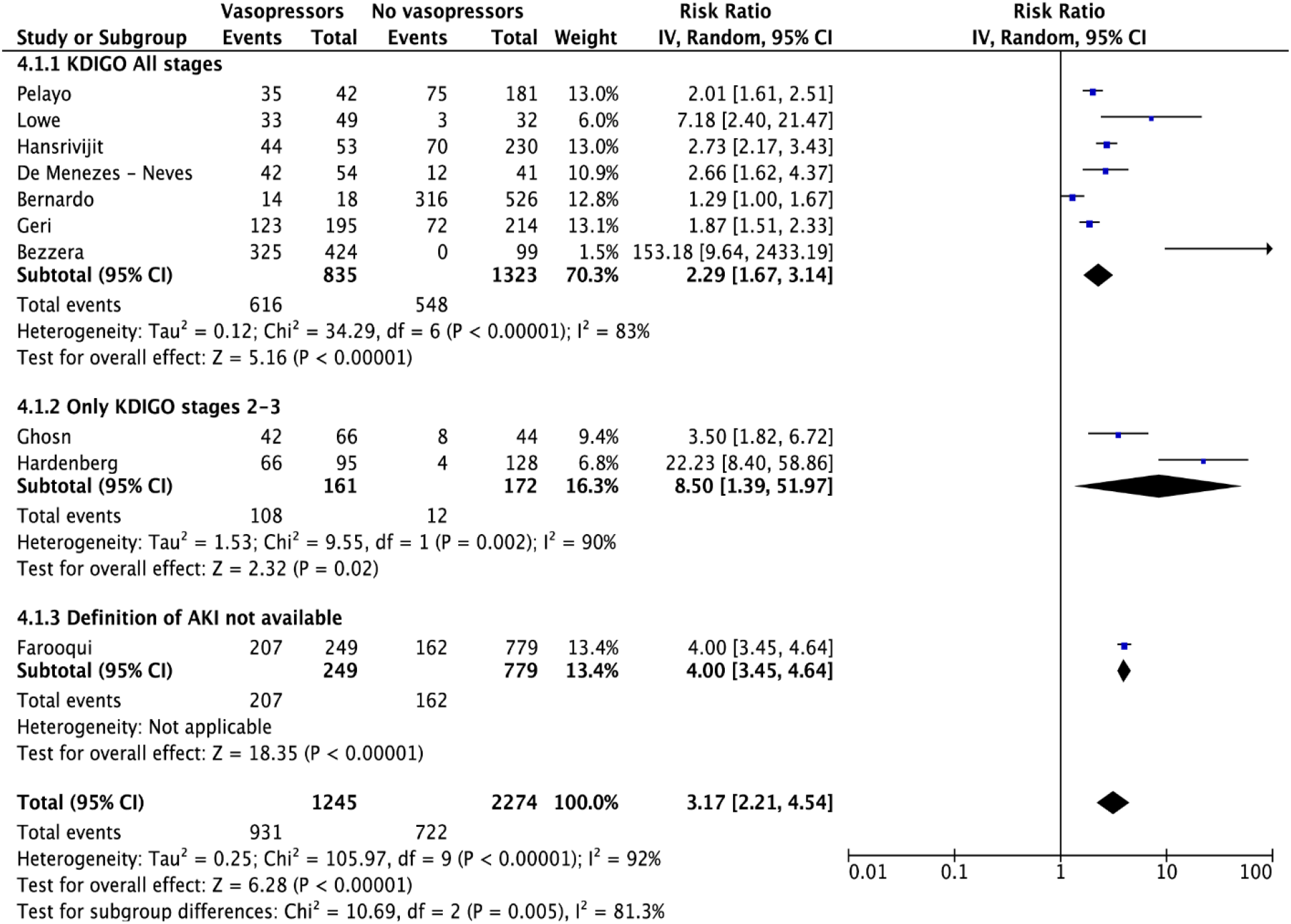
Subgroup analyses were performed based on the definition of acute kidney injury

##### Other secondary outcomes

No data were identified for the remaining secondary outcomes.

### Risk of bias, quality of evidence

The overall quality of the studies, as assessed by the MINORS tool, ranged between moderate and high. The exact score for each study is available in Appendix C3. In addition, visual inspection of the funnel plot (Appendix D2) and the Begg’s test (p=0.18) did not reveal significant publication bias for the studies included in the AKI analysis. All included studies were observational non-randomized studies with striking heterogeneity that does not allow to derive an estimate of overall effect. According to GRADE criteria, the quality of evidence provided by the studies was low.

## DISCUSSION

Many high-quality RCTs have addressed the effects of vasopressors on the outcomes of non-covid patients, yet their impact on mortality in patients with COVID-19 had not been studied so far. The most important finding of this systematic review and meta-analysis is that vasopressor use is associated with mortality in critically ill patients with COVID-19. Although these results are based on published non-randomized evidence, they raise significant concerns for the routine management of these individuals.

The mortality of critically ill patients with COVID-19 is high [53, 54]. A main cause is the characteristics of the SARS-CoV-2 infection, which can rapidly affect other organs including the cardiovascular system [55]. Although administration of vasopressors is a fundamental treatment of hypotension, the traditional (non-covid) hemodynamic management of shock and the adverse effects of vasoactive agents may be associated with complications and poor outcome in patients with COVID-19. Indeed, the present analysis reveals an association between vasopressor use and in-hospital mortality or mortality at 30 days. This association may be coincidental due to the severity of critical illness. However, norepinephrine, the suggested first-line agent (weak recommendation) [56], have numerous biological effects that can affect many systems including the immune and hematological systems, the renin-angiotensin-aldosterone system, the cardiovascular and respiratory systems, and others [13], suggesting a complex interplay that can have a detrimental effect on survival. Future RCTs should investigate the degree to which the association between vasopressor use and mortality is causal.

Of the 33 included studies in this systematic review and meta-analysis, only four studies included data on specific vasopressors. The highest mortality rate was observed in patients receiving vasopressin or epinephrine. Although one can appraise that these patients had severe shock necessitating second- and third-line vasopressors [7], these observations merit further discussion. Epinephrine is well-known for its adverse effects in non-covid and COVID-19 patients [57], but our observations regarding vasopressin are quite interesting. Although vasopressin infusion reduces total norepinephrine-equivalent dose requirements and may be renal and pulmonary vasculature sparing [58], there is evidence showing a pronounced activation of the vasopressin system in COVID-19 patients and that molecular complexes form between the SARS-CoV-2 spike protein, soluble angiotensin-converting enzyme-2 (ACE2), and vasopressin, facilitating cellular infection and aggravating outcome [59, 60]. However, data from a small clinical cohort did not show a clinically relevant effect of vasopressin infusion on viral mRNA level in critically ill patients with COVID-19 (but who were not treated with corticosteroids or interleukin-6 antagonists) [58]. The findings of the present analysis suggest that the addition of arginine vasopressin agonists might not be a good choice for these individuals. Considering that vasopressin is suggested as a second-line vasopressor in the latest international guidelines [7, 56], further research is required to establish the therapeutic efficiency of vasopressin in critically ill patients with COVID-19.

Another intriguing finding is the low mortality rate in patients receiving angiotensin-II as a sole or second-line vasopressor agent. Serpa Neto et al. showed a potentially positive effect of angiotensin-II on blood pressure and fraction of inspired oxygen in COVID-19 patients, but they did not collect data regarding treatment with steroids or other drugs, which may have affected their results [49]. Leisman et al. and Ofosu-Barko et al. reported that angiotensin-II treatment was associated with rapid improvement in multiple physiologic indices [38, 51]. The rationale for angiotensin-II therapy is based on decreasing the expression of the ACE2 receptors, which can reduce the entry of the COVID-19 virus into cells [61, 62]. However, the progressive loss of ACE2 shifts the system to an overall higher angiotensin level due to the impaired ability of ACE2 to degrade it, which may explain the initial hemodynamic stability of patients with COVID-19 [63]. Therefore, exogenous use of angiotensin-II may be harmful in patients with increased endogenous levels. Taking into consideration its potential favorable effects in critically ill patients with COVID-19, randomized controlled trials are needed to further evaluate angiotensin-II for the treatment of COVID-19-related shock.

A meta-analysis of randomized clinical trials with non-covid patients reported that vasopressor therapy is not associated with differences in mortality in the overall population, while prophylactic administration in patients with vasodilatory shock may improve survival [64]. In addition, a Cochrane systematic review found no evidence of substantial differences in total mortality between several vasopressors [65]. Nevertheless, vasopressors are a heterogeneous class of drugs with powerful and immediate haemodynamic effects, and each drug has advantages and disadvantages. These characteristics are particularly important in patients with COVID-19 who are characterized by unique pathophysiological disturbances and different hemodynamic phenotypes that necessitate a thorough understanding of the underlying complex pathophysiology and careful selection and administration of vasoactive agents.

In COVID-19, the progressive hypoxemia initially increases cardiac output and capillary recruitment, which maintain microcirculatory oxygen-extraction capacity by increasing red blood cell availability (silent hypoxia) [66–68]. However, microcirculatory flow decreases proportionally to the increasing inflammation, hypercoagulation, and thrombosis, resulting in multi-organ failure at later stages [66,69–71]. In the study by Mesquida et al., patients showed alterations in systemic microcirculatory status, and the degree of these alterations correlated with the severity of the respiratory disease [24]. The relationship between MAP and organ blood flow may be different in critically ill patients with COVID-19 and improving only macrocirculation might be inadequate to maintain tissue perfusion. In these patients, vasopressor use can overwhelm endogenous receptor-mediated vessel regulation, further contributing to the loss of hemodynamic coherence [67, 72], and therefore, hemodynamic management should focus on optimizing microcirculatory perfusion instead of attaining a predefined MAP target.

### Limitations

This meta-analysis was based on observational studies. Due to the lack of randomized controlled trials, the synthesis of all the available knowledge on the specific outcomes was difficult. This is an inherent problem to observational studies, and not least considering the difficulties of collecting data during the periods of surges across the globe. Moreover, we could not obtain individual data to reach a minimal level of evidence that could result on relevant findings and thus, the effects of different vasopressors may reflect differences in severity and/or practices. International registries are necessary to collect uniform data to evaluate the effect of vasopressors on mortality and other outcomes in critically ill patients with COVID-19.

In addition, the level of heterogeneity was high and the conclusions drawn from this review must be cautious and reserved. This analysis included patients from various settings, i.e., HDU, ICU, and Emergency Department. Consequently, it may have included heterogeneous groups of patients with COVID-19. No data from the included studies was available to adjust the resulting odds ratios according to age, comorbidities, the presence of septic shock, or other known factors that affect ICU mortality. Also, most of the secondary outcomes could not be assessed. Another limitation is the heterogeneity of definitions of AKI that were used across different studies. Finally, non-English publications were not included.

## CONCLUSIONS

Vasopressor use was associated with an increase in in-hospital mortality, 30-day mortality, and acute kidney injury in critically ill patients with COVID-19. The lower mortality rate in patients receiving angiotensin-II as a sole or second-line vasopressor agent and the higher mortality rate in patients receiving vasopressin and epinephrine are worth noting. However, the included studies were observational non-randomized studies with striking heterogeneity that does not allow to derive an estimate of overall effect. Randomized controlled trials are required to estimate the correlation of specific vasopressors with adverse effects and mortality in this population.

## PERSPECTIVES

The results of the present systematic review and meta-analysis suggest for early administration of low-dose vasopressors, with or without inodilator agents, in an effort to avoid excessive doses that could have detrimental effect on survival, especially at later disease stages. An alternative second-line vasopressor may be angiotensin-II. However, further immediate research is recommended to elucidate the effects of angiotensin-II and other vasopressors acting through pathways other than the adrenergic pathway as sole or second-line vasopressor agents. These agents may be associated with a significant increase in survival.

A possible explanation for the association of vasopressors with mortality may lie in the microcirculation [66–72]. The physiological pulsatile shear stress from normal laminar flow maintains the normal endothelial cell functions and the expression of ACE2s and other anticoagulant/antithrombotic and antioxidant substances [73]. However, dysfunctional endothelium resulting from turbulent flow displays a hypercoagulant/prothrombotic and pro-oxidant state and impairs microcirculatory reactivity and flow [74]. Therapeutic approaches should consider the systemic vascular involvement, allowing an individualized, physiology-guided management. It is almost certain that there are distinct COVID-19 phenotypes/subphenotypes that include the impairment of microvasculature as key feature and their identification will have important therapeutic implications [75]. Of note, an increased CVP in critically ill patients with COVID-19 may impair venous return and retrogradely increase post-capillary venular pressure which, together with excessive vasopressor doses, impair capillary perfusion and increase the diffusion distance of oxygen [76, 77]. Thus, minimizing fluid administration is also crucial for improving tissue perfusion in this population.

The present systematic review and meta-analysis analyzed data from retrospective studies only. Further research and well-designed trials are necessary to investigate the effect of the type (catecholamines *vs.* non-catecholamines), time of administration, and infusion rates of vasopressors in order to develop more specific treatment strategies and integrate a more individualized approach in patients with COVID-19. Although designing and conducting RCTs on vasopressors during a disease outbreak may be difficult, the need for assessing their effect on the outcome of critically ill patients with COVID-19 is imperative. We recommend the exploration of suitable animal models and the integration of translational research to aid in the identification of the most suitable vasopressor in this population and also, to better define homogenous target (sub)populations for trials [78–80]. At the same time, large pragmatic generalizable RCTs with very broad inclusion criteria will help improve generalizability of the findings.

## Data Availability

Data can be made available upon request after publication through a collaborative process. Researchers should provide a methodically sound proposal with specific objectives in an approval proposal. Please contact the corresponding author for additional information.

## Acknowledgements

We would like to thank Dr. George L. Anesi, University of Pennsylvania Perelman School of Medicine, for responding to our request for additional study data.

## Author contributions

Conceptualization: AC. Data curation: MM, GM. Formal analysis: MM, GM. Methodology: AC, MM, GM, EL, NP, IP. Project administration: AC. Visualization: MM, GM, EL, NP, IP, AC. Writing -original draft: AC. Writing - review & editing: AC, MM, GM, EL, NP, IP.

## APPENDIX LEGENDS

**Appendix A:** PRISMA checklist.

**Appendix B:** Algorithms used for all databases.

**Appendix C1:** Definitions used for AKI and mortality follow-up time points.

**Appendix C2:** Angiotensin-related data.

**Appendix C3:** MINORS results for each study.

**Appendix D1:** Funnel plot for mortality meta-analysis.

**Appendix D2:** Funnel plot for AKI meta-analysis.

